# Physiology-guided cardioneuroablation for vagally mediated syncope and functional bradyarrhythmias

**DOI:** 10.64898/2026.08.03.26359633

**Authors:** Gurukripa N. Kowlgi, J. C. Pachón-M, Narut Prasitlumkum, Arvind Gulati, Jeanwoo Yoo, Kallie Karlen, Nicholas Y. Tan, Alan M. Sugrue, Ammar M. Killu, Abhishek J. Deshmukh, Suraj Kapa, Christopher V. DeSimone, Freddy Del-Carpio Munoz, Konstantinos C. Siontis, Malini Madhavan, Peter A. Noseworthy, Paul A. Friedman, E. I. Pachón-M, Carlos Thiene C. Pachón, Juan Carlos Zerpa, Yong-Mei Cha, Win-Kuang Shen, Samuel J. Asirvatham

## Abstract

**Background:** Vagally mediated syncope and functional bradyarrhythmias can cause recurrent symptoms, injury and a need for permanent pacing, yet standard therapies do not address the underlying autonomic reflex. Cardioneuroablation (CNA) targets this mechanism, but adoption has been limited by uncertainty about patient selection, procedural endpoints and durable outcomes.

**Methods:** We studied 95 consecutive patients with vagally mediated syncope (n=78) or functional bradyarrhythmia (n=17) who underwent CNA at a single center by a single lead operator under a uniform, prospectively maintained protocol. Extracardiac vagal stimulation (ECVS) was performed before and after ablation to confirm vagally mediated sinus or atrioventricular (AV) nodal responses and to verify their attenuation. Outcomes through 1 year included clinical recurrence, quality of life, pacing burden and pacemaker extraction.

**Results:** Before ablation, ECVS provoked sinus pauses in 95.8% of patients and AV block in 91.6%; after ablation, sinus pauses were abolished in every patient and residual AV block was present in 4.2%. One-year freedom from recurrent syncope and from bradyarrhythmia-related events was 93.8% and 94.1%. Syncope burden fell from a median of 2.7 episodes per year to none, and disease-specific quality of life improved substantially (P<0.001). Among patients with pre-existing devices, atrial pacing burden fell from a median of 22% to 0%. There were no strokes, deaths or myocardial infarctions.

**Conclusions:** In this single-center cohort, physiology-guided CNA was associated with durable symptom control, improved quality of life and reduced pacing across vagally mediated bradyarrhythmia syndromes. Multicenter controlled studies are needed to confirm these findings.

## Introduction

Vagally mediated syncope and functional bradyarrhythmias are common, and most patients do well with reassurance and conservative measures. A subset, however, experience recurrent symptoms, injury, impaired quality of life or pacing requirement despite standard care^1–3^. Permanent pacing can reduce recurrent syncope in carefully selected patients with documented vagal slowing of the heart rate or atrioventricular (AV) conduction, but it does not modify the underlying autonomic reflex and may be undesirable in younger patients or in those without intrinsic conduction system disease^1–4^. Cardioneuroablation (CNA) has therefore emerged as a catheter-based strategy to attenuate excessive parasympathetic influence on sinoatrial (SA) and AV nodal function^5^.

Although CNA was first described more than two decades ago, its adoption has remained limited by uncertainty about patient selection, procedural technique and outcome assessment^3,5,6^. Referring clinicians are often unsure which patients with recurrent syncope or functional bradyarrhythmias to send for ablation. For electrophysiologists, published approaches differ in mapping strategy, ablation targets, procedural endpoints and follow-up measures^3,7^. Recent studies, including systematic reviews, randomized trials and multicenter registries, suggest that CNA can reduce recurrent syncope in selected patients^8–12^. However, outcomes vary across studies, likely reflecting differences in patient selection, ablation strategy and procedural endpoints^8–14^. A recent single-center study further emphasized this issue by showing that recurrent syncope after CNA was common when procedural approaches were heterogeneous, but that durable response was associated with achieving multiple intraprocedural physiologic markers of autonomic modification^13^. These uncertainties have made it difficult to translate CNA into a universally accepted referral and treatment pathway.

Extracardiac vagal stimulation (ECVS) has been previously described as a method to provoke and reassess parasympathetic effects on sinus and AV nodal function during CNA^6,15^. Whether this physiologic evidence of autonomic modification translates into durable clinical benefit in a larger, carefully phenotyped cohort has been less clear. In this prospectively maintained single-center registry, we evaluated patients undergoing CNA for vagally mediated syncope or functional bradyarrhythmias. We focused on detailed phenotyping, ECVS-guided assessment of autonomic modification, and follow-up measures that included symptom recurrence, heart rate, heart rate variability, quality of life, pacing burden and pacemaker system extraction.

## Methods

### Study design and oversight

We analyzed consecutive patients with vagally mediated syncope or functional bradyarrhythmias who underwent cardioneuroablation (CNA) at Mayo Clinic between July 2022 and February 2026 and were enrolled in a prospectively maintained institutional registry. Referral triage data were summarized separately for patients referred or triaged for possible CNA evaluation between July 2022 and December 2025. The study was approved by the Mayo Clinic Institutional Review Board. Patients provided research authorization according to institutional requirements. The present analysis was restricted to patients with at least 3 months of clinical follow-up. Patients undergoing CNA primarily for atrial fibrillation were not included in this analysis.

### Study population

Patients were considered for CNA when symptoms were attributed to excessive vagal influence on SA node or AV nodal function and when conservative management was ineffective, not tolerated or unlikely to provide durable benefit. Clinical evaluation included review of symptom history, syncope burden, medication use, electrocardiography (ECG), ambulatory rhythm monitoring, implantable loop recorder (ILR) data when available, pacemaker interrogation when applicable and head-up tilt-table testing (HUTT) when performed. Patients with evidence of intrinsic sinus node disease, structural conduction system disease or another primary explanation for symptoms were not considered appropriate candidates for CNA. Baseline characteristics are summarized in Table 1.

**Table 1.** Baseline characteristics by clinical phenotype.

| Characteristic | Overall cohort<br>(n = 95) | Syncope-predominant<br>(n = 78) | Functional bradyarrhythmia-predominant<br>(n = 17) |
| --- | --- | --- | --- |
| **Demographics and comorbidities** |  |  |  |
| Age at CNA, years | 40 [32-58] | 37.5 [32-55.8] | 58 [33-69] |
| Female sex | 48 (50.5) | 43 (55.1) | 5 (29.4) |
| BMI, kg/m<sup>2</sup> | 26.4 [23.4-30.8] | 26.3 [23.3-30.3] | 26.6 [24.2-32.4] |
| White race/ethnicity | 90 (94.7) | 73 (93.6) | 17 (100.0) |
| LVEF, % | 61 [58.5-63] | 61 [59-63] | 61 [58-63] |
| Hypertension | 30 (31.6) | 20 (25.6) | 10 (58.8) |
| Diabetes mellitus | 9 (9.5) | 7 (9.0) | 2 (11.8) |
| Coronary artery disease | 12 (12.6) | 9 (11.5) | 3 (17.6) |
| Heart failure with reduced ejection fraction | 2 (2.1) | 2 (2.6) | 0 (0.0) |
| Prior atrial fibrillation | 21 (22.1) | 15 (19.2) | 6 (35.3) |
| Inappropriate sinus tachycardia | 9 (9.5) | 9 (11.5) | 0 (0.0) |
| Postural orthostatic tachycardia syndrome | 6 (6.3) | 5 (6.4) | 1 (5.9) |
| Obstructive sleep apnea | 13 (13.7) | 9 (11.5) | 4 (23.5) |
| **Clinical presentation** |  |  |  |
| Syncope/presyncope indication | 78 (82.1) | 78 (100.0) | 0 (0.0) |
| Functional bradycardia indication | 36 (37.9) | 20 (25.6) | 16 (94.1) |
| AV block indication | 19 (20.0) | 17 (21.8) | 2 (11.8) |
| Syncope-related trauma | 25 (26.3) | 25 (32.1) | 0 (0.0) |
| Syncopal episodes before CNA, median [Q1-Q3] |  | 9 [3-15] | N/A |
| Patients with known syncope episode count, n | 75 | 75 | 0 |
| Prior medical therapy tried | 33 (34.7) | 29 (37.2) | 4 (23.5) |
| **Documented bradyarrhythmia phenotype** |  |  |  |
| Sinus bradycardia alone | 76 (80.0) | 61 (78.2) | 15 (88.2) |
| AV block alone | 4 (4.2) | 4 (5.1) | 0 (0.0) |
| Sinus bradycardia and AV block | 15 (15.8) | 13 (16.7) | 2 (11.8) |
| **Tilt-table testing** |  |  |  |
| Tilt-table report available | 80 (84.2) | 71 (91.0) | 9 (52.9) |
| Positive tilt-table response | 41/80 (51.2) | 40/71 (56.3) | 1/9 (11.1) |
| Mixed response among positive tests | 16/41 (39.0) | 16/40 (40.0) | 0/1 (0.0) |
| Cardioinhibitory response among positive tests | 19/41 (46.3) | 19/40 (47.5) | 0/1 (0.0) |
| Vasodepressor response among positive tests | 6/41 (14.6) | 5/40 (12.5) | 1/1 (100.0) |
| Asystole >3 s on tilt-table testing | 22/80 (27.5) | 22/71 (31.0) | 0/9 (0.0) |

Values are median [Q1-Q3], n (%) or n/N (%). Percentages use the column denominator unless a row explicitly provides n/N.
Positive tilt-table response was defined as mixed, cardioinhibitory or vasodepressor response among patients with a tilt-table report available.
Phenotype assignment was based on the complete clinical evaluation and not on tilt-table findings alone.
AV = atrioventricular; BMI = body mass index; CNA = cardioneuroablation; LVEF = left ventricular ejection fraction.

### Clinical assessment and phenotype classification

Patients were classified into two clinical phenotypes before ablation. The syncope-predominant phenotype included patients with recurrent reflex or vagally mediated syncope, with or without documented sinus pauses, AV block or mixed vasodepressor features. The functional bradyarrhythmia phenotype included patients with symptomatic sinus bradycardia, sinus pauses or functional AV block in the absence of recurrent syncope and without evidence of intrinsic conduction system disease.

Tilt-table responses, when available, were classified according to the dominant clinical and hemodynamic pattern. Cardioinhibitory responses were defined by prominent vagal slowing of sinus rate or atrioventricular conduction. Vasodepressor responses were defined by predominant hypotension without a major bradycardic component. Mixed responses included both hypotension and bradycardia. Final phenotype assignment was based on the totality of clinical, rhythm-monitoring and tilt-table data rather than tilt-table findings alone.

### Cardioneuroablation procedure

All procedures were performed under general anesthesia. Electroanatomic mapping was performed using a contemporary three-dimensional mapping system. Ablation targeted atrial ganglionated plexus regions implicated in parasympathetic control of sinus and atrioventricular nodal function. Target selection was based on anatomic location, electrogram characteristics and responses to extracardiac vagal stimulation. Lesion delivery was individualized according to the presenting phenotype and intraprocedural response, with additional ablation performed when residual vagally mediated sinus node or atrioventricular nodal responses persisted.

Ablation could be performed in the left atrium, right atrium or both, depending on the clinical phenotype and residual response to stimulation. Core regions included the right superior ganglionated plexus region and the left atrial ridge or ligament of Marshall region, with additional ganglionated plexus sites targeted as needed. Radiofrequency settings, catheter type, mapping system and procedural details were recorded prospectively in the registry.

### Extracardiac vagal stimulation

Extracardiac vagal stimulation was performed before and after ablation to assess parasympathetic effects on SA and AV nodal function, as previously described^11^. Stimulation was delivered with a dedicated high-frequency vagal stimulation system at 50 Hz, 80 mA and 0.2-ms pulse width for 5 seconds. Baseline responses included sinus slowing or pause, AV block and duration of the longest sinus pause. After ablation, stimulation was repeated to assess residual vagal effects. The procedural endpoint was abolition or marked attenuation of vagally mediated SA node and AV nodal responses. Additional markers included an increase in resting sinus rate and improvement in AV nodal conduction parameters when applicable.

### Follow-up and outcomes

Patients were followed through outpatient visits, electronic medical record review, rhythm monitoring, ILR data and pacemaker interrogation when available. The primary outcome for the syncope-predominant phenotype was recurrent syncope after cardioneuroablation. The primary outcome for the functional bradyarrhythmia phenotype was recurrent symptomatic bradyarrhythmia or permanent pacemaker implantation after CNA.

Secondary outcomes included changes in heart rate, heart rate variability, AV nodal conduction, pacing burden, pacemaker system extraction and disease-specific quality of life. In patients with pre-existing pacemakers, atrial and ventricular pacing burdens were compared before and after ablation. Pacemaker extraction was recorded when performed after clinical and device-based assessment. Quality of life was assessed using the Impact of Syncope on Quality of Life (ISQL) questionnaire when available, with lower scores indicating better quality of life. When post-CNA tilt-table testing was performed, responses were classified using the same categories as baseline testing. Paired tilt-table analyses were restricted to patients with both pre- and post-CNA tilt-table results available.

Safety outcomes included vascular complications, pericardial effusion or hemopericardium, pericarditis, gastroparesis, inappropriate sinus tachycardia, new atrial arrhythmias, ventricular arrhythmias, stroke, myocardial infarction, phrenic nerve injury, urinary retention and death. Adverse events were adjudicated by review of the medical record.

### Statistical analysis

Continuous variables are reported as mean ± standard deviation or median [Q1–Q3], according to distribution. Categorical variables are reported as counts and percentages. Paired pre- and post-ablation variables were compared using paired t tests or Wilcoxon signed-rank tests, as appropriate. Paired binary tilt-table components were compared using exact McNemar tests. Categorical variables were compared using chi-square or Fisher exact tests. Time-to-event outcomes were estimated using the Kaplan–Meier method. Given the small number of recurrent clinical events, multivariable modeling was not performed. All tests were 2-sided, and a P value <0.05 was considered statistically significant. Statistical analyses were performed using R (R Foundation for Statistical Computing, Vienna, Austria), and figures were generated using Python (Python Software Foundation, Wilmington, DE).

## Results

### Study cohort and clinical phenotypes

Between July 2022 and December 2025, approximately 350 patients were referred or triaged for possible CNA evaluation. Of these, 130 were offered CNA consultation and 95 underwent CNA after clinical evaluation, testing and shared decision-making. Patients not offered consultation were generally redirected because of low likelihood of benefit, insufficient evidence for a vagally mediated mechanism or an alternative diagnosis (Figure S1). The final cohort included 78 patients with syncope-predominant disease and 17 patients with functional bradyarrhythmia- predominant disease (Fig. 1 and Table 1). Median age was 40 years [32-58], 48 patients (50.5%) were female and median left ventricular ejection fraction was 61% [58.5-63]. Patients with syncope-predominant disease were younger than those with functional bradyarrhythmia-predominant disease, and hypertension was more common in the functional bradyarrhythmia group. Syncope-related trauma occurred in 25 patients (26.3%), all in the syncope-predominant group. Among patients with known syncope episode count, the median number of lifetime syncopal episodes before CNA was 9 [3-15]. Tilt-table reports were available in 80 patients (84.2%), of whom 41 (51.2%) had a positive response, although clinical phenotype was not determined by tilt-table findings alone.

**Figure 1.**
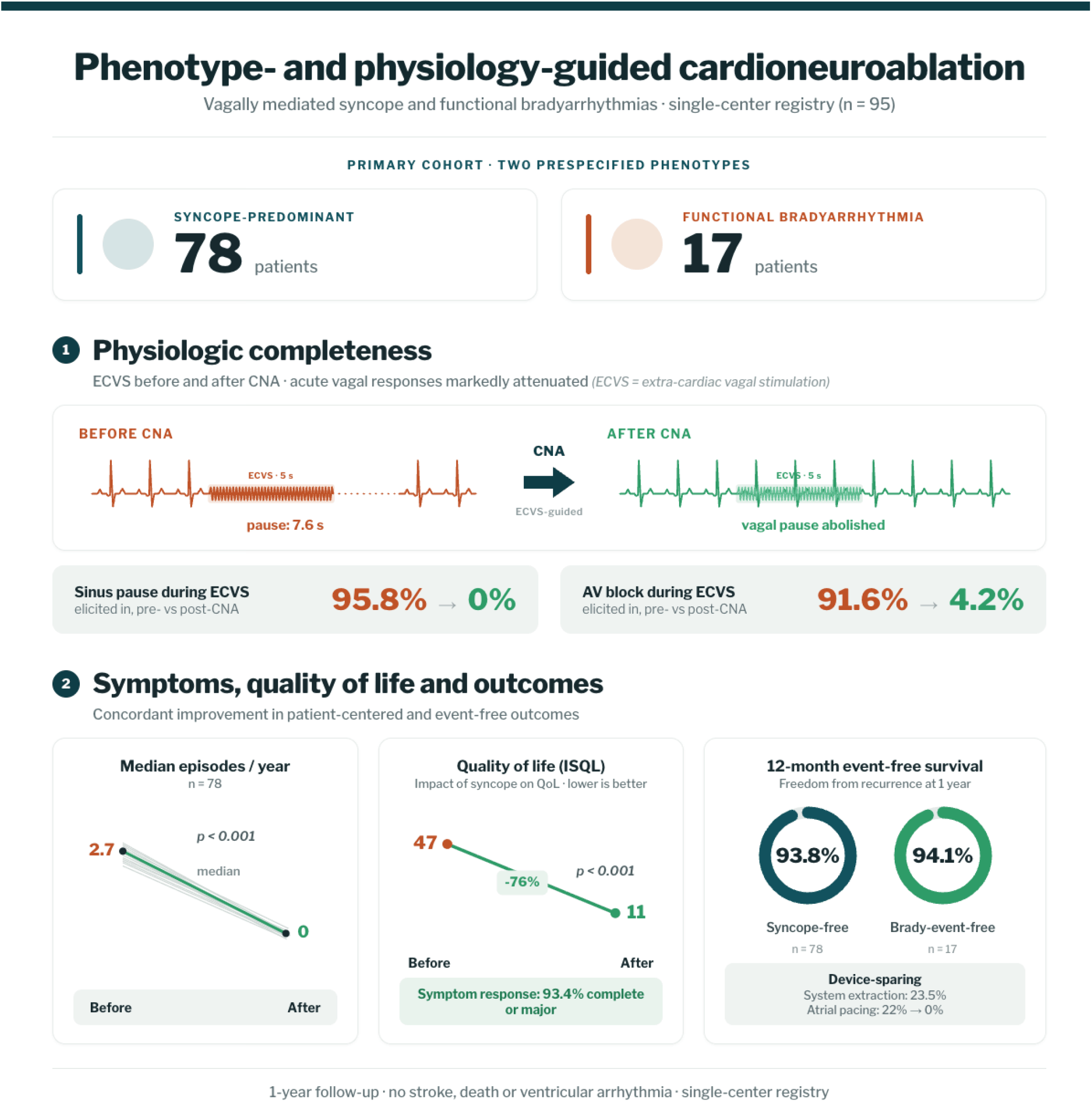
Phenotype- and physiology-guided cardioneuroablation for vagally mediated syncope and functional bradyarrhythmias. Central summary of the study design and principal findings. Ninety-five patients underwent cardioneuroablation (CNA) for one of two prespecified clinical phenotypes: syncope-predominant disease or functional bradyarrhythmia-predominant disease. Extracardiac vagal stimulation (ECVS) was performed before and after ablation to assess vagally mediated effects on sinus and atrioventricular nodal function. After CNA, ECVS-provoked sinus pauses were abolished, and residual atrioventricular block was uncommon. Clinical follow-up demonstrated reduction in syncope burden, improvement in disease-specific quality of life, high 1-year freedom from recurrent syncope or bradyarrhythmia-related events, and reduced pacing burden among patients with pre-existing pacemakers. ISQL denotes Impact of Syncope on Quality of Life.

### Procedural characteristics and acute physiologic response

All patients underwent CNA under general anesthesia with electroanatomic mapping (Table S1). Ablation targeted prespecified ganglionated plexus regions, including the Marshall tract or left atrial appendage ridge in all patients, the right superior ganglionated plexus in all patients, the aortocaval ganglionated plexus in 88 patients (92.6%), the left atrial posteromedial ganglionated plexus in 73 patients (76.8%) and the right atrial posteromedial ganglionated plexus in 80 patients (84.2%).

Before ablation, ECVS produced a sinus pause in 91 patients (95.8%) and AV block in 87 patients (91.6%). The median longest sinus pause during pre-ablation ECVS was 7.65 seconds [5.0-9.75]. Repeat ECVS was performed after ablation in 93 patients (97.9%). After CNA, ECVS-provoked sinus pause was abolished in all patients, and residual AV block was present in 4 patients (4.2%) (Fig. 1 and Table S1).

### Clinical outcomes through 1 year

Through 1 year of follow-up, recurrent syncope occurred in 6 of 78 patients with syncope- predominant disease. The estimated 1-year freedom from recurrent syncope was 93.8% (Fig. 2a). Among patients with recurrent syncope, only one had recurrent cardioinhibitory syncope and underwent repeat CNA. The remaining recurrent events did not have clear recurrent cardioinhibitory physiology documented on available follow-up evaluation.

**Figure 2.**
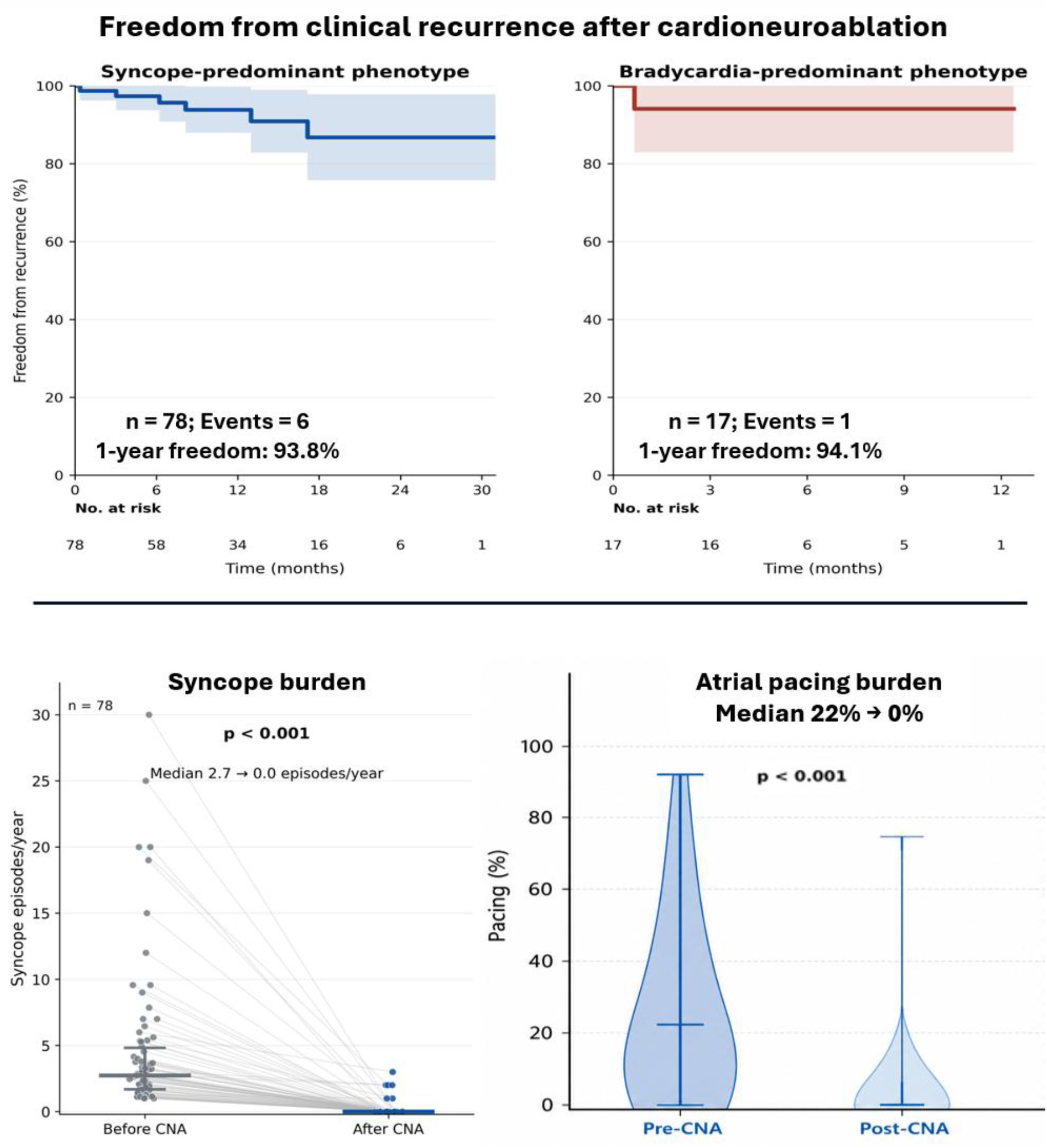
Clinical recurrence, syncope burden and pacing burden after cardioneuroablation. Kaplan-Meier estimates of freedom from clinical recurrence are shown for the syncope-predominant phenotype (a) and the functional bradyarrhythmia-predominant phenotype (b). In the syncope-predominant group, the primary outcome was recurrent syncope after cardioneuroablation (CNA). In the functional bradyarrhythmia-predominant group, the primary outcome was recurrent symptomatic bradyarrhythmia or permanent pacemaker implantation after CNA. Shaded areas represent 95% confidence intervals, and numbers at risk are shown below each plot. Syncope burden before and after CNA is shown among patients with syncope-predominant disease (c). Atrial pacing burden before and after CNA is shown among patients with pre-existing pacemakers (d). P values were calculated using paired tests.

In the functional bradyarrhythmia-predominant group, 1 of 17 patients developed recurrent symptomatic bradyarrhythmia requiring permanent pacemaker implantation. The estimated 1- year freedom from bradyarrhythmia-related events was 94.1% (Fig. 2b). Among patients with syncope-predominant disease, syncope burden decreased from a median of 2.7 episodes per year before CNA to 0 episodes per year after CNA (P < 0.001) (Fig. 2c). Among the 17 patients with pre-existing pacemakers, atrial pacing burden decreased from a median of 22% before CNA to 0% after CNA (P < 0.001) (Fig. 2d). Of these, four patients (23.5%) underwent complete pacemaker system extraction after CNA.

### Autonomic, conduction and hemodynamic changes

CNA was associated with changes consistent with reduced parasympathetic influence on sinus and AV nodal function (Fig. 3, Figure S2 and Table 2). Intraprocedural heart rate increased from 59.1 ± 14.1 bpm before ablation to 81.1 ± 14.8 bpm after ablation (P < 0.001). Atrioventricular Wenckebach cycle length shortened from 550 ms [400-600] to 400 ms [345-430] (P < 0.001), whereas the HV interval did not change significantly. Systolic blood pressure decreased modestly, whereas diastolic blood pressure did not change significantly.

**Figure 3.**
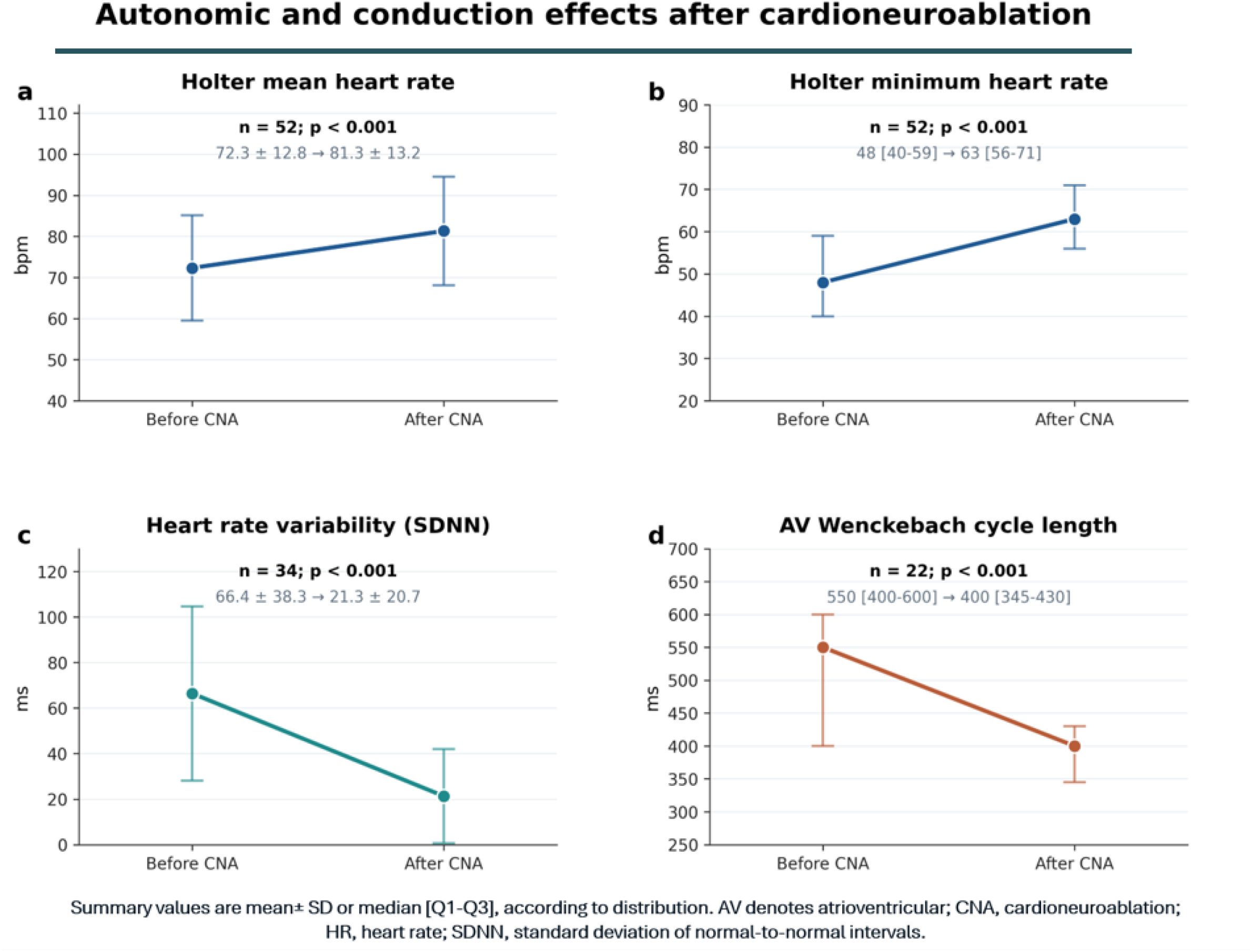
Autonomic and conduction changes after cardioneuroablation. Summary physiologic measures before and after cardioneuroablation (CNA) are shown for ambulatory heart rate, Holter minimum heart rate, heart rate variability and atrioventricular nodal conduction. Holter mean heart rate and Holter minimum heart rate increased after CNA, whereas heart rate variability decreased and atrioventricular Wenckebach cycle length shortened. Values are shown as mean ± SD or median [Q1-Q3], according to distribution. P values were calculated using paired tests. Patient-level paired data are shown in Figure S2. AV denotes atrioventricular; CL, cycle length; HR, heart rate; SDNN, standard deviation of normal-to-normal intervals.

**Table 2.** Autonomic, conduction and hemodynamic changes after CNA.

| Variable | Before CNA | After CNA | P value |
| --- | --- | --- | --- |
| **Intraprocedural parameters** |  |  |  |
| EP lab heart rate, bpm | 59.1 ± 14.1 | 81.1 ± 14.8 | **<0.001** |
| AV Wenckebach cycle length, ms | 550 [400-600] | 400 [345-430] | **<0.001** |
| HV interval, ms | 48 [41.5-52] | 47.5 [42-51] | 0.78 |
| **Hemodynamic Parameters** |  |  |  |
| Systolic BP (mmHg) | 123.6 ± 16.7 | 119.4 ± 16.2 | **0.033** |
| Diastolic BP (mmHg) | 75.3 ± 9.0 | 73.7 ± 9.9 | 0.19 |
| **Three-month ECG and ambulatory parameters** |  |  |  |
| ECG PR interval, ms | 166.2 ± 31.8 | 157.9 ± 25.6 | **0.032** |
| QT interval, ms | 407 [382-434] | 376 [362.5-394] | **<0.001** |
| Holter minimum heart rate, bpm | 48 [40-59] | 63 [56-71] | **<0.001** |
| Holter mean HR (bpm) | 72.3 ± 12.8 | 81.3 ± 13.2 | **<0.001** |
| Holter maximum heart rate, bpm | 123.8 ± 29.5 | 122.8 ± 22.4 | 0.83 |
| SDNN, ms | 66.4 ± 38.3 | 21.3 ± 20.7 | **<0.001** |
| pNN50, % | 32.5 [20-68.5] | 5.0 [0-8.75] | **<0.001** |

**Abbreviations:** AV, atrioventricular; CNA, cardioneuroablation; ECG, electrocardiogram; EP, electrophysiology; HR, heart rate; HV, His-ventricular; pNN50, percentage of adjacent normal-to-normal intervals differing by more than 50 ms; SDNN, standard deviation of normal-to-normal intervals. Values are mean ± SD or median [Q1-Q3]

At 3 months, electrocardiographic PR interval shortened from 166.2 ± 31.8 ms to 157.9 ± 25.6 ms (P = 0.032), and QT interval shortened from 407 ms [382-434] to 376 ms [362.5-394] (P < 0.001). Holter minimum heart rate increased from 48 bpm [40-59] to 63 bpm [56-71] (P < 0.001), and Holter mean heart rate increased from 72.3 ± 12.8 bpm to 81.3 ± 13.2 bpm (P < 0.001). Holter maximum heart rate did not change significantly. Heart rate variability decreased, with SDNN decreasing from 66.4 ± 38.3 ms to 21.3 ± 20.7 ms (P < 0.001) and pNN50 decreasing from 32.5% [20-68.5] to 5.0% [0-8.75] (P < 0.001).

Among the subset of 65 patients with paired pre- and post-CNA tilt-table testing, positive tilt responses decreased after ablation (Table S2). This paired cohort differs from the full baseline tilt-table cohort summarized in Table 1. Before CNA, 30 of 65 patients (46.2%) in the paired cohort had a positive tilt response, including 10 mixed, 6 vasodepressor and 14 cardioinhibitory responses. After CNA, 52 patients (80.0%) had a negative tilt test, 11 (16.9%) had a vasodepressor response, 2 (3.1%) had a cardioinhibitory response and none had a mixed response. The cardioinhibitory component (pure cardioinhibitory plus mixed responses) decreased from 24 of 65 patients (36.9%) to 2 of 65 patients (3.1%; P < 0.001), whereas the vasodepressor component did not change significantly. Among patients with baseline mixed or vasodepressor responses in the paired cohort, 10 of 16 (62.5%) became tilt-negative after CNA.

### Quality of life, pacing burden and patient-reported response

Disease-specific quality of life improved substantially after CNA (Fig. 4a). ISQL score decreased from 47 [41-50] before CNA to 11 [8-16] after CNA (P < 0.001), indicating improved quality of life. Among 91 patients with available clinical response data, 51 (56.0%) reported complete symptom resolution, 34 (37.4%) reported major improvement, 3 (3.3%) reported partial improvement and 3 (3.3%) reported no improvement (Fig. 4b). Overall, 85 of 91 patients (93.4%) reported either complete symptom resolution or major improvement.

**Figure 4.**
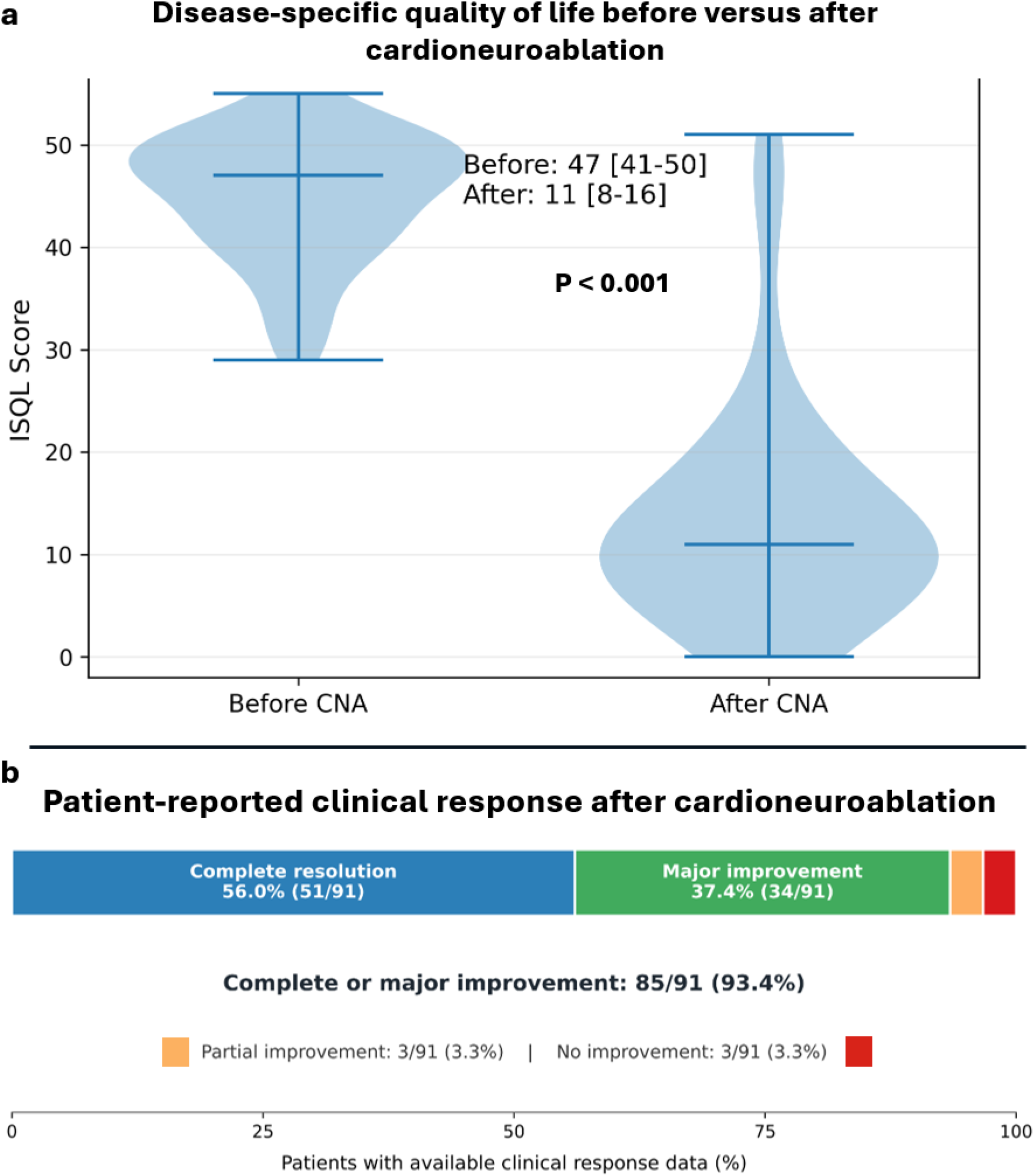
Quality of life and patient-reported clinical response after cardioneuroablation. Disease-specific quality of life before and after cardioneuroablation (CNA) was assessed using the Impact of Syncope on Quality of Life (ISQL) questionnaire, with lower scores indicating better quality of life (a). Patient-reported clinical response after CNA is shown as complete resolution, major improvement, partial improvement or no improvement among patients with available response data (b). The ISQL analysis included patients with paired pre- and post-CNA questionnaire data. Categorical response assessment included 91 patients with available clinical response data. P value was calculated using the Wilcoxon signed-rank test.

### Safety

Safety outcomes are summarized in Table 3. Vascular complications occurred in 2 patients (2.1%), hemopericardium in 1 patient (1.0%), pericarditis in 7 patients (7.3%) and gastroparesis in 3 patients (3.1%). Headache occurred in 9 patients (9.4%) and resolved within 1-2 weeks. Inappropriate sinus tachycardia was present in 13 patients (13.7%) at 3 months and persisted in 3 patients (3.1%) at 6 months; 1 patient (1.1%) remained on treatment for persistent inappropriate sinus tachycardia at last follow-up. There were no strokes, myocardial infarctions, deaths or phrenic nerve injuries. No new ventricular arrhythmias were observed. There were also no new atrial arrhythmias or urinary retention.

**Table 3.** Safety outcomes after CNA.

| **Periprocedural complications** | **N (%)** |
| --- | --- |
| Vascular (Hematoma/Pseudoaneurysm/ AV fistula) | 2 (2.1) |
| Pericarditis | **7 (7.3)\*** |
| Hemopericardium | 1(1.1) |
| Phrenic nerve injury | 0 |
| Stroke | 0 |
| Myocardial infarction | 0 |
| Death | 0 |
| **CNA-specific/post-procedural effects** |  |
| Headaches | **9 (9.4)\*** |
| IST at 3 months | **13(13.7)** |
| IST at 6 months | **3(3.1)** |
| IST at 12 months/Last follow-up requiring Rx | **1 (1.1)** |
| New atrial arrhythmias | 0 |
| New Ventricular arrhythmias (PVC/VT) | 0 |
| Gastroparesis | **3 (3.1)** |
| Urinary retention | 0 |
| *Values are n (%). Percentages use the full cohort denominator of 95 patients. Headache resolved within 1-2 weeks after ablation. IST at 3,6, and 12 months refers to inappropriate sinus tachycardia documented during follow-up. Persistent IST requiring treatment at last follow-up was present in 1 patient. AV, atrioventricular; CNA, cardioneuroablation; IST, inappropriate sinus tachycardia; PVC, premature ventricular contraction; VT, ventricular tachycardia.* |  |
| *\*Resolved within 1-2 weeks post-ablation* |  |

## Discussion

In this prospectively maintained single-center registry, CNA for vagally mediated syncope and functional bradyarrhythmias was associated with consistent acute attenuation of vagal responses, low rates of recurrent clinical events, improved disease-specific quality of life and reduced pacing burden. Although CNA and ECVS have been previously described, this study extends prior work by applying a systematic phenotype-based and physiology-guided workflow in a rigorously characterized cohort with longitudinal clinical, pacing-related and patient-reported outcomes^3,5,6^. These findings are clinically relevant because uncertainty regarding patient selection, procedural endpoints and expected outcomes has limited broader adoption of CNA despite more than two decades of experience^3,5,7^.

A practical contribution of this study is the use of a structured clinical pathway for a problem that remains incompletely defined in current practice. Current syncope guidelines define diagnostic evaluation and pacing indications, but not referral criteria for CNA^8,10,11,13^. In this cohort, candidacy was based on the combined assessment of clinical phenotype, rhythm documentation, tilt-table findings, and device interrogation when available. At our center, this structured assessment has helped identify patients likely to benefit from CNA. We present the resulting pathway (Fig. 5) as the approach used in the present cohort rather than as a formal recommendation for other center.

**Figure 5.**
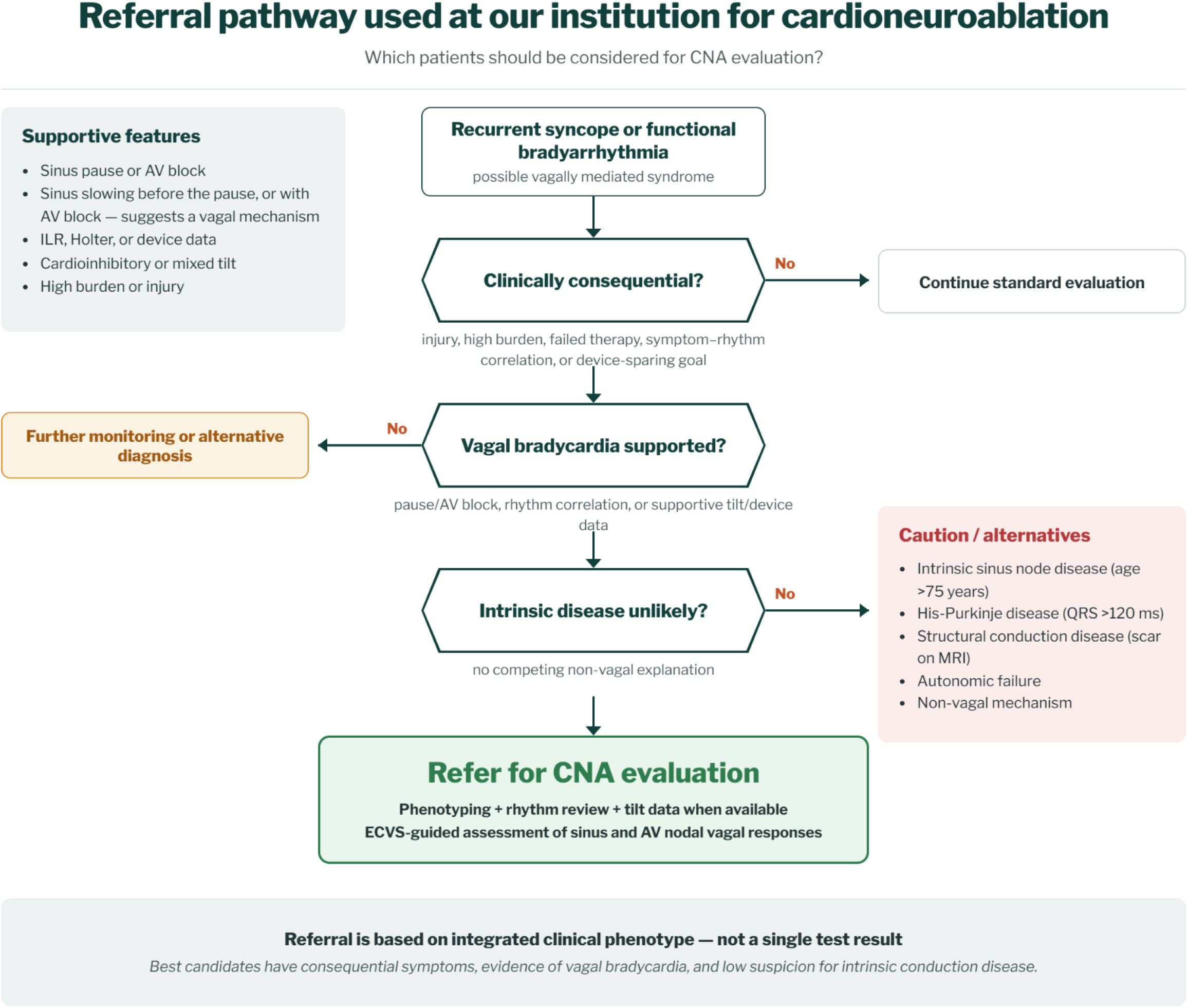
Referral pathway used at our institution for cardioneuroablation evaluation. Referral pathway used at our institution to identify patients considered for cardioneuroablation (CNA) evaluation, shown as the approach applied in the present cohort rather than as a general recommendation. At our center, patients with recurrent syncope or functional bradyarrhythmia were assessed for features supporting a vagally mediated mechanism, including a classic vasovagal pattern, documented sinus pause or atrioventricular block, symptom-rhythm correlation, cardioinhibitory or mixed tilt-table response, high symptom burden, syncope-related injury or lifestyle limitation. Patients were considered most appropriate for referral when symptoms were clinically consequential, vagal bradycardia was supported by clinical, rhythm, device or tilt-table data, and intrinsic conduction system disease or competing non-vagal causes were unlikely. Features that should prompt caution or alternative evaluation include intrinsic sinus node disease, His-Purkinje disease, structural conduction disease, autonomic failure, primary orthostatic hypotension or another non-vagal mechanism. AV denotes atrioventricular; ECVS, extracardiac vagal stimulation; ILR, implantable loop recorder.

The present findings should also be interpreted in light of a recent study evaluating physiologic markers of effective autonomic denervation during CNA for vasovagal syncope. In that cohort, recurrent syncope occurred in 44% of patients during intermediate-term follow-up, but recurrence was substantially lower among patients who achieved multiple favorable intraprocedural markers, including vagal response during left-sided ganglionated plexus ablation, sinus rate acceleration and absence of atropine responsiveness^9,11,12,14^. Those findings suggest that clinical durability depends on the degree of autonomic modification achieved, rather than on the anatomic lesion set alone. We build on that concept by evaluating a larger cohort that included both syncope-predominant and functional bradyarrhythmia-predominant phenotypes, with integrated clinical, physiologic, quality-of-life and pacing-related outcomes.

ECVS was central to the physiologic strategy used in this cohort. It has been previously described as a method to provoke and reassess parasympathetic effects during ablation procedures^11,12,14^. In the present cohort, nearly all patients had marked vagal responses before ablation, whereas sinus pauses were abolished and residual AV block was uncommon after CNA. These findings support ECVS as a practical intraprocedural assay of the autonomic target, complementing anatomic and electrogram-based ablation strategies. At the same time, ECVS should be interpreted as an acute physiologic endpoint rather than proof of durable neural elimination, which makes longitudinal clinical follow-up essential^3,6^.

Clinical outcomes in this cohort were favorable in the context of prior randomized and registry experience. Recurrent syncope occurred in a small proportion of patients with syncope- predominant disease, and bradyarrhythmia-related events were uncommon in patients with functional bradyarrhythmia-predominant disease. These findings are broadly consistent with prior studies showing clinical benefit after CNA in selected patients with reflex syncope, functional atrioventricular block and functional bradycardia^11,12,14^. The event rates and complication profile in the present cohort also compare well with contemporary multicenter experience, although these comparisons should be interpreted cautiously because of differences in patient selection, procedural strategy, follow-up duration and recurrence definitions^16^.

The recurrence pattern also supports the importance of careful phenotyping. In an exploratory comparison of syncope-predominant patients with and without recurrent syncope, no baseline physiologic or procedural variable clearly distinguished patients with recurrence, although patients with recurrence had higher baseline ISQL scores and the small number of events limits interpretation (Table S3). The fact that most recurrent events were not clearly cardioinhibitory on available follow-up evaluation suggests that residual vasodepressor susceptibility, mixed reflex physiology or noncardiac mechanisms may contribute to symptoms after CNA.

The paired tilt-table findings further clarify what physiology CNA appears to modify. CNA is designed primarily to attenuate excessive parasympathetic effects on sinus and AV nodal function, not to eliminate all causes of hypotension or transient loss of consciousness. Consistent with this mechanism, the cardioinhibitory component of tilt response decreased substantially after CNA, whereas the vasodepressor component did not change significantly. Among patients with baseline mixed or vasodepressor tilt responses, 62.5% (10 of 16) became tilt-negative after CNA, and 25% (4 of 16) had residual vasodepressor responses. These findings suggest that CNA may reduce the bradycardic component of reflex syncope even when vasodepressor physiology is present, but persistent hypotensive susceptibility may still contribute to recurrent symptoms. The inclusion of selected patients with mixed or vasodepressor features reflects real-world referral practice and supports careful phenotyping rather than rigid exclusion based on a single tilt-table pattern^1–3^.

The improvement in quality of life and reduction in pacing burden are important because the clinical value of CNA is not limited to preventing recurrent syncope. Recurrent vagally mediated symptoms can impair daily function even when events are not frequent enough to meet conventional pacing thresholds. In this cohort, disease-specific quality-of-life scores improved substantially, more than 90% of patients reported complete symptom resolution or major improvement, atrial pacing burden decreased among patients with pre-existing pacemakers and nearly one quarter underwent complete pacemaker system extraction after clinical and device-based assessment. Prior work has reported discontinuation of permanent pacing and pacemaker extraction after CNA in patients with vagally mediated bradycardia, supporting the concept that pacing therapy may not always be lifelong when the dominant mechanism is autonomic rather than intrinsic conduction system disease^3,12^. The present findings extend those observations by evaluating pacing-related outcomes within a larger, carefully phenotyped cohort that also incorporated systematic ECVS-guided physiologic assessment, symptom follow-up and disease-specific quality-of-life assessment^3,5,16^.

The physiologic changes observed after CNA were consistent with targeted parasympathetic attenuation rather than nonspecific autonomic disruption. Resting heart rate increased, AV nodal conduction improved and heart rate variability decreased, while Holter maximum heart rate did not change significantly. Systolic blood pressure decreased modestly, whereas diastolic blood pressure did not change significantly. This pattern suggests that CNA modified vagal effects on sinus and AV nodal function without producing a generalized hypertensive response or an increase in maximum ambulatory heart rate. Safety outcomes were also favorable, with low rates of serious complications and no stroke, death, new ventricular arrhythmia, or urinary retention. These findings compare favorably with contemporary multicenter CNA experience, although differences in patient selection, ablation strategy and follow-up duration limit direct comparison^3,12^.

This study has limitations. Although the registry was prospectively maintained, the present analysis was observational, single-center and nonrandomized, and the absence of a sham or medically treated control group limits causal inference. Patient selection was based on detailed clinical evaluation at an experienced center, which may limit generalizability. Follow-up duration was modest, and longer follow-up is needed to define durability, late recurrence and the stability of autonomic modification. Tilt-table testing, rhythm monitoring, device interrogation and quality-of-life assessment were not uniformly available in all patients, and paired tilt-table analysis was restricted to patients with both pre- and post-CNA tilt testing. Finally, the small number of recurrent clinical events limited the ability to identify predictors of recurrence, and recurrent symptoms after CNA may reflect heterogeneous mechanisms, including residual vasodepressor susceptibility, mixed reflex physiology or noncardiac causes.

In summary, CNA was associated with favorable clinical, physiologic and patient-reported outcomes in selected patients with vagally mediated syncope and functional bradyarrhythmias. By combining rigorous clinical phenotyping, systematic ECVS-guided physiologic assessment and longitudinal follow-up, this study supports a practical framework for identifying and treating patients who may benefit from CNA. Multicenter controlled studies are needed to confirm these findings, refine referral criteria and define the durability of this approach.

### Clinical Perspective

#### What Is New?

- In a single-center cohort of 95 patients treated with a uniform, physiology-guided cardioneuroablation protocol, extracardiac vagal stimulation performed before and after ablation confirmed acute attenuation of vagally mediated sinus and atrioventricular nodal responses in nearly all patients.
- Through 1 year, the approach was associated with high freedom from recurrent syncope and bradyarrhythmia-related events, substantial improvement in disease-specific quality of life, and a marked reduction in atrial pacing burden, including pacemaker extraction in a subset.
- Paired pre- and post-ablation tilt-table testing showed that the cardioinhibitory component of the reflex was reduced whereas the vasodepressor component was not, clarifying which physiology cardioneuroablation modifies.

#### What Are the Clinical Implications?

- Cardioneuroablation guided by intraprocedural physiologic confirmation may offer a mechanism-targeted, pacing-sparing option for selected patients with vagally mediated syncope or functional bradyarrhythmia, many of them young.
- Because most affected patients first present to primary care and general medicine, these data are relevant to referral decisions beyond the electrophysiology laboratory.
- Multicenter, controlled studies are needed to confirm durability, refine patient selection and define the role of cardioneuroablation relative to pacing and conservative care.

### Use of large language model

During manuscript preparation, the authors used Anthropic Claude as an editorial review tool to assist with language refinement, organization, consistency checks and figure layout concepts. All scientific content, analyses, data interpretation, figures and conclusions were developed, verified and approved by the authors. No large language model was used to generate primary data, perform unsupervised data analysis or make scientific conclusions.

## Data Availability

Deidentified data supporting the findings of this study may be made available from the corresponding author upon reasonable request and after approval by the Mayo Clinic Institutional Review Board and completion of applicable data use agreements.

## Author Contributions

G.N.K. conceived and designed the study, curated and analyzed the data, performed the procedures, and wrote the original draft. A.G., J.Y., G.N.K. and K.K. contributed to data collection and registry management. G.N.K., N.P., N.Y.T., A.M.S., A.M.K., A.J.D., S.K., C.V.D., F.D.-C.M., K.C.S., M.M., P.A.N., Y.-M.C., W.-K.S., P.A.F. and S.J.A. contributed to patient care, and data interpretation. J.C.P.-M., E.I.P.-M., C.T.C.P. and J.C.Z. contributed the extracardiac vagal stimulation methodology and procedural expertise. All authors interpreted the data, critically revised the manuscript for important intellectual content and approved the final version. P.A.F. and S.J.A. supervised the work.

## Sources of Funding

This study received no specific funding.

## Disclosures

J.C.P.-M., E.I.P.-M. and C.T.C.P. are inventors on intellectual property related to a dedicated vagal stimulation device used for extracardiac vagal stimulation. S.J.A. is an inventor on devices and methods for neurocardiac modulation, which were not used in this study, and is a speaker and consultant for Abbott, Boston Scientific, Medtronic, Biotronik and Johnson & Johnson. The remaining authors declare no competing interests.

## Funding Sources

None

## Disclosures

J.C.P.-M., E.I.P.-M. and C.T.C.P. are inventors on intellectual property related to a dedicated vagal stimulation device used for extracardiac vagal stimulation. The device was used in preclinical studies related to this work and in a subset of human procedures. S.J.A. is an inventor on devices and methods for neurocardiac modulation, which were not used in this study; he is also a speaker and consultant for Abbott, Boston Scientific, Medtronic, Biotronik and Johnson & Johnson. No funding or resources from these relationships were used for this study. The remaining authors declare no competing interests relevant to this manuscript.

## Abbreviations

AV: atrioventricular
BP: blood pressure
CNA: cardioneuroablation
ECG: electrocardiogram
ECVS: extracardiac vagal stimulation
HR: heart rate
HRV: heart rate variability
HUTT: head-up tilt-table testing
ILR: implantable loop recorder
ISQL: Impact of Syncope on Quality of Life questionnaire
IST: inappropriate sinus tachycardia
QOL: quality of life
SA: sinoatrial

## Notes

### Author Declarations

The study was approved by the Mayo Clinic Institutional Review Board. Patients provided research authorization according to institutional requirements.

## References

1. Shen W-K, Sheldon RS, Benditt DG, Cohen MI, Forman DE, Goldberger ZD, Grubb BP, Hamdan MH, Krahn AD, Link MS, Olshansky B, Raj SR, Sandhu RK, Sorajja D, Sun BC, Yancy CW. 2017 ACC/AHA/HRS guideline for the evaluation and management of patients with syncope: A report of the American college of cardiology/American heart association task force on clinical practice guidelines and the heart rhythm society. Circulation. 2017;136:e60–e122.

2. Brignole M, Moya A, de Lange FJ, Deharo J-C, Elliott PM, Fanciulli A, Fedorowski A, Furlan R, Kenny RA, Martiın A, Probst V, Reed MJ, Rice CP, Sutton R, Ungar A, van Dijk JG. 2018 ESC Guidelines for the diagnosis and management of syncope. Kardiol Pol. 2018;76:1119–1198.

3. Aksu T, Brignole M, Calo L, Debruyne P, Biase LD, Deharo JC, Fanciulli A, Fedorowski A, Kulakowski P, Morillo C, Moya A, Piotrowski R, Stec S, Sutton R, van Dijk JG, Wichterle D, Tse HF, Yao Y, Sheldon RS, Vaseghi M, Pachon JC, Scanavacca M, Meyer C, Amin R, Gupta D, Magnano M, Malik V, Schauerte P, Shen WK, Carlos Zerpa Acosta J. Cardioneuroablation for the treatment of reflex syncope and functional bradyarrhythmias: A Scientific Statement of the European Heart Rhythm Association (EHRA) of the ESC, the Heart Rhythm Society (HRS), the Asia Pacific Heart Rhythm Society (APHRS) and the Latin American Heart Rhythm Society (LAHRS). Europace [Internet]. 2024;26. Available from: 10.1093/europace/euae206

4. Kapa S, Venkatachalam KL, Asirvatham SJ. The autonomic nervous system in cardiac electrophysiology: an elegant interaction and emerging concepts. Cardiol Rev. 2010;18:275–284.

5. Pachon JC, Pachon EI, Cunha Pachon MZ. “Cardioneuroablation” – new treatment for neurocardiogenic syncope, functional AV block and sinus dysfunction using catheter RF-ablation. Europace. 2005;7:1–13.

6. Pachon M. JC, Pachon M. EI, Santillana P. TG, Lobo TJ, Pachon CTC, Pachon M. JC, Albornoz V. RN, Zerpa A. JC. Simplified Method for Vagal Effect Evaluation in Cardiac Ablation and Electrophysiological Procedures. JACC Clin Electrophysiol. 2015;1:451–460.

7. Penela D, Berruezo A, Roten L, Futyma P, Richter S, Chun J. Cardioneuroablation for vasovagal syncope. Results from an EHRA Survey. Europace. 2024;26:euae102.290.

8. Vandenberk B, Lei LY, Ballantyne B, Vickers D, Liang Z, Sheldon RS, Chew DS, Aksu T, Raj SR, Morillo CA. Cardioneuroablation for vasovagal syncope: A systematic review and meta-analysis. Heart Rhythm. 2022;19:1804–1812.

9. Piotrowski R, Baran J. Cardioneuroablation for reflex syncope: Randomized controlled trial. JACC Clin Electrophysiol. 2023;9:1–12.

10. Tu B, Chen A, Cai S, Zhang Z, Zhou L, Lai Z, Maimaitijiang P, Hu Z, Wu L, Ding L, Zheng L, Yao Y. The efficacy of left atrial vs biatrial cardioneuroablation in patients with Vasovagal Syncope: A randomized clinical trial. JACC Clin Electrophysiol. 2025;11:1265– 1276.

11. Aksu T, Piotrowski R, Tung R, De Potter T, Markman TM, du Fay de Lavallaz J, Rekvava R, Alyesh D, Joza JE, Badertscher P, Do DH, Bradfield JS, Upadhyay G, Sood N, Sharma PS, Guler TE, Gul EE, Kumar V, Koektuerk B, Dal Forno ARJ, Woods CE, Rav-Acha M, Valeriano C, Enriquez A, Sundaram S, Glikson M, D’Avila A, Shivkumar K, Kulakowski P, Huang HD. Procedural and Intermediate-term Results of the Electroanatomical-guided Cardioneuroablation for the Treatment of Supra-Hisian Second- or Advanced-degree Atrioventricular Block: the PIRECNA multicentre registry. Europace [Internet]. 2024;26. Available from: 10.1093/europace/euae164

12. Tung R, Pujol-Lopez M, Locke AH, Alyesh DM, Sundaram S, Shah AD, Kumar V, Kowlgi G, Kumar K, Shvilkin A, Aksu T, Vasaiwala S, Weiss JP, Zawaneh M, Winterfield JR, John LA, Santangeli P, Woods C, Tzou WS, Kapur S, Sauer W, Thosani AJ, Dewland TA, Gerstenfeld EP, Upadhyay GA, d’Avila A. Cardioneural ablation for functional bradycardia and vasovagal syncope: Outcomes from the U.s. multicenter CNA registry. JACC Clin Electrophysiol. 2025;11:1683–1695.

13. Vojnika J, Patel D, Enriquiez A, Hyman MC, Dixit S, Santangeli P, Nazarian S, J Callans D, Frankel DS, E Marchlinski F, Markman TM. Physiological markers of effective autonomic denervation are associated with outcomes after cardioneuroablation for vasovagal syncope. JACC Clin Electrophysiol [Internet]. 2026;Available from: 10.1016/j.jacep.2026.05.021

14. Gigante C, Penela D, Viveros D, Falasconi G, Teresi L, Latini AC, Soto-Iglesias D, Franco- Ocaña P, Francia P, Alderete J, Turturiello D, Bellido AF, Zaraket F, Valeriano C, Mea R, Tonello B, Sanchez-Mollá L, De Lucia C, Matiello M, Fernández-Armenta J, San Antonio R, Saglietto A, Ortiz-Pérez J-T, Marti-Almor J, Berruezo A. A Tailored Approach to Cardioneuroablation for Reflex Syncope and Functional Bradycardia. Results from the ELEGANCE multicenter study. Europace [Internet]. 2025;28. Available from: 10.1093/europace/euaf320

15. Steiger K, Kashou AH, Ledet CB, Kowlgi GG. Successful cardioneural ablation for deglutition syncope. Heart Lung. 2026;78:102819.

16. Stec S, Wileczek A, Reichert A, Śledź J, Kosior J, Jagielski D, Polewczyk A, Zając M, Kutarski A, Karbarz D, Zyśko D, Nowarski Ł, Stodółkiewicz-Nowarska E. Shared decision making and cardioneuroablation allow discontinuation of permanent pacing in patients with vagally mediated bradycardia. J Cardiovasc Dev Dis. 2023;10:392.

